# Seroprevalence of SARS-CoV-2 in Palestine: a cross-sectional seroepidemiological study

**DOI:** 10.1101/2020.08.28.20180083

**Authors:** Nouar Qutob, Faisal Awartani, Zaidoun Salah, Mohammad Asia, Imad Abu Khader, Khaled Herzallah, Nadeen Balqis, Husam Sallam

## Abstract

Seroprevalence rates are important indicators to the epidemiology of COVID-19 and the extent of the pandemic given the existence of asymptomatic cases. The purpose of this study is to assess the seroprevalence rate in the Palestinian population residing in the West Bank. Blood samples were collected between 15^th^ June 2020 and 30^th^ June 2020 from 1355 individuals from randomly selected households in the West Bank in addition to 1136 individuals visiting Palestinian medical laboratories between the 1^st^ May 2020 and 9^th^ July 2020 for a routine checkup. Out of the 2491 blood samples collected, serological tests for 2455 adequate serum samples were done using an Immunoassay for qualitative detection of antibodies against SARS-CoV-2. The random sample of Palestinians living in the West Bank yielded 0% seroprevalence with 95% CI [0,0.0036], while the lab referrals sample yielded an estimated seroprevalence of 0.354% with 95% CI [0.0011,0096]. Our results indicate that as of July 2020, seroprevalence in Palestine persist low and is inadequate to provide herd immunity, emphasizing the need to maintain health measures to keep the outbreak under control. Population-based seroprevalence studies are to be conducted periodically to monitor the SARS-CoV-2 seroprevalence in Palestine and inform policy makers about the efficacy of their surveillance system.

## Introduction

Coronavirus disease 2019, known as COVID-19, is an infectious respiratory disease caused by novel coronavirus SARS-CoV-2^1^. Since its emergence in Wuhan, China in December 2019^2^ SARS-CoV-2 has spread rapidly around the globe, ultimately being declared by the World Health Organization (WHO) as a global pandemic ^3,4^ and new cases and deaths are being reported daily^5^.

Most authorities rely on PCR testing results to estimate number of COVID-19 cases and make up-to-date decisions ^6^. Thus, numbers of patients tested positive for SARS-CoV-2 through PCR testing, symptomatic patients, those admitted to hospitals, or deceased from Cobvid-19 are updated on a daily basis. However, the data may exclude a fraction of the population with previous mild or asymptomatic COVID-19that has not been tested by PCR. The proportion of the population who have overcome the infection without being noticed can probably be approximated by testing for antibodies against SARS-CoV-2. Antibodies may confer immunity to repeat infection and a high proportion of immune individuals can attenuate the epidemic. Measures of anti-SARS-CoV-2 seroprevalence can also be used to estimate the clinical impact of COVID-19. To this effect, several serological surveys of SARS-CoV-2 have been done worldwide ^7–16^.

There is lack of data on the the percentage of undiagnosed Palestinian population with previous mild or asymptomatic COVID-19. Prevalence of COVID-19 among Palestinian remains unknown and many are concerned about this uncertainty. To this end, we conducted a population cross sectional based seroepidemiological study to assess the spread of SARS-CoV-2 throughout the West Bank. The study included 2491 individuals, designed to be representative by cities (1355 from randomly selected households and 1136 from laboratory referrals). Elecsys® Anti-SARS-CoV-2 testing was done on 2455 adequate serum samples. Here, we describe the study design and the results of the first wave of the study.

## Methods

### Study design and participants

The study conducted is a cross-sectional serologic testing study aimed to investigate seropositivity for SARS-CoV-2 in the non-institutionalized Palestinian population residing in the West Bank.

The study involved 1355 participants from 11 governorates, including 112 localities (supplementary table 1). 1395 households were selected using 3-stage cluster sampling. The cluster of households or census track is considered to be a geographic location that is comprised of approximately 100 households. The process for conducting cluster sampling was carried as follows: (1) Selecting a cluster of households, (2) Selecting 10 households randomly from each cluster and (3) Selecting a person at random from the selected household. The clusters were selected using probability proportional to size (PPS) sampling (Table 1).

**Table 1:** The PPS sampling algorithm

| Location | # of households in the location | Cumulative |
| --- | --- | --- |
| L1 | X1 | C1=X1 |
| L2 | X2 | C2=X1+X2 |
| L3 | X3 | C3=X1+X2+X3 |
| . | . | . |
| . | . | . |
| . | . | . |
| Lk-1 | Xk-1 | Ck-1=X1+X2+...+Xk-1 |
| Lk | Xk | Ck = X1+X2+...+Xk |

To select the number of clusters within each population location: (1) we calculated the sampling interval which equals the total number of households divided by the total number of clusters need to be selected by the sample say for example (m). So the sampling interval SI= N/m, where N is the total number of households. (2) selected a random number R0 between 0 and SI. (3) calculated Ri as R0+i*SI, a cluster is selected in Li if Ri belongs to the interval [Ci-1, Ci].

Field work was carried out between 15th^th^ June 2020 and 30^th^ June 2020 by a team of registered nurses, laboratory technicians, nursing students and laboratory technician students from the Arab American University following standardized health protocols (World Health, 2020).

The study also included 1136 participants from Medicare laboratory referrals between 1^st^ May 2020 and 9^th^ July 2020 in 16 branches in the West Bank (supplementary table 2).

Participants donated a blood sample for antibodies detection. Blood samples were centrifuged and serum was separated, labelled, stored at −20C at AAUP laboratory until it was used.

Approval from National ethical committee was obtained (PHRC/HC/737/20). Written informed consent was obtained from the 1355 study participants and approvals were obtained from Medicare laboratories for samples to be tested.

### Detection of antibodies

Serological tests for 2455 adequate serum samples were done using an Immunoassay for the qualitative detection of antibodies against SARS-CoV-2 (Elecsys® Anti-SARS-CoV-2) in human serum by using the Cobas analyzer cobas e 411 (Roche). The assay uses a recombinant protein representing the nucleocapsid (N) antigen for the determination of antibodies against SARS-CoV-2 with testing time of 18 minutes. We included 6 samples from recovered cases with detected SARS-COV2 antibodies as a positive control.

The manufacturers reported clinical specificity of 99.81 % (99.65 – 99.91 %) using a total of 5,272 samples (from diagnostic routine, blood donors, a common cold panel, and a coronavirus panel*) obtained before December 2019 and a clinical sensitivity of 65.5 % (56.1 – 74.1 %) 0-6 days post PCR confirmation, 88.1 % (77.1 – 95.1 %) 7-13 days post PCR and 100 % (88.1 – 100 %) >14 days post PCR confirmation. This was done using total of 204 samples from 69 symptomatic patients with a PCR confirmed SARS-CoV-2 infection. One or more consecutive specimens from these patients were collected after PCR confirmation at various time points.

### Statistical Analysis

We estimated seroprevalence as the proportion of individuals who had a positive result in the total SARS-CoV-2 antibodies in the immunoassay.

We used Wilson Method With Continuity Correction and Boundary Truncation (WCCBT) to construct 95% CI for the population parameter of seroprevalence ^17^.

## Results

Of the 1395 eligible individuals residing in households selected using 3-stage cluster sampling, 1355 participants agreed to participate in the study. The proportion of females was lower compared to males (137, 1218 respectively) including 349 in age group (15-24), 314 in age group (25-34), 377 in age group (35-49) and 315 in age group (50+). Out of the 1355 blood samples collected, 1319 serum samples were adequate for testing. None of the tested specimens revealed presence of antibodies against SARS-CoV-2. A 95% CI for the population parameter of seroprevalence was [0,0.0036].

Of the1136 participants from Medicare laboratory referrals in 16 branches. The proportion of males was lower than females (395, 741 respectively) including 71 in age group lower less than 15, 173 in age group (15-24), 297 tests in age group (25-34), 290 in age group (35-49) and 305 in age group (50+). All serum samples were adequate for testing. Out of the 1136 tested participants, 3 males, ages 38, 58, 59 and 1 female, age 40 revealed antibodies against Sars-CoV-2 with 95% CI [0.0011,0096].

## Discussion

To our knowledge, this is the first SARS-CoV-2 seroprevalence study in Palestine. The findings from this seroprevalence study for SARS-CoV-2 indicate that the estimated seroprevalence of the total SARS-COV2 antibodies persist low. The random sample of Palestinians living in the West Bank yeilded 0% seroprevalence with 95% CI [0,0.0036], while the lab referrals sample yielded an estimated seroprevalence of 0.354% with 95% CI [0.0011,0096]. Seroprevelance in Palestine is very close to that identified in Jordan (0% prevalence)^15^, a neighboring country, which was explained to be due to the strict closures implemented by the Jordanian government and the eventual curtailing of infections in Jordan. In comparison to other countries like Spain, Italy. Japan India, Los Angeles, Germany, Switzerland the seroprevalence in Palestine is low ^7–16^. It is, however, noteworthy that a comparison with other countries may be problematic due to the timing and the stage of the pandemic which may vary affecting the seroprevalence estimates.

A key strength of our study is the random selection of households residents collected between 15^th^ June 2020 and 30^th^ June 2020. The clusters were selected using probability proportional to size (PPS) sampling. This technique ensures getting self-weighting sample which can be used to produce unbiased estimators for the parameters of interest. However, samples from female were more difficult to obtain due to a cultural inhibition regarding allowing nurses to enter the households. Also, random sample did not include children under the age of 15 to avoid anxiety due to fear of needles in the study to eliminate personal fears due to blood withdrawl.

As for the lab referals sample, it represents the population of the lab referals between the 1^st^ May 2020 and 9^th^ July 2020. The seroprevalence in this sample was 4 positive cases out of the 1136 samples tested. Since the seroprevalence within the lab referral population is close to zero, we used Wislon Method With Continuity Correction and Truncation (WCCBT) to construct a 95% confidence intervale for the population parameter of seroprevalence [0.0011,0096].

Our study only detected Sars-Cov2 antibodies. However, it is important to recognize that cellular immunity may play a role in providing immunity against SARS-CoV-2 reinfection^18^. Further studies aimed at testing cellular immunity are important. It is also noteworthy that previous studies have indicated that asymptomatic individuals were reported to have a weaker immune response to SARS-CoV-2 infection and a higher percentage of asymptomatic individuals became seronegative when compared to symptomatic individuals in the early recovering phases. The reduction in neutralizing antibody levels may have implications for immunity strategy and serological surveys ^19^.

In conclusion, our study provides estimates of SARS-CoV-2 seroprevalence in Palestine. Our estimate is low indicating that as of July 2020 the population does not have herd immunity. In this situation, health measures have to be taken to keep the outbreak under control. In order to monitor the SARS-CoV-2 seroprevalence in Palestine and inform policy makers about the efficacy of their survalance system, conducting population-based seroprevalence studies on a regular basis is important.

## Data Availability

Data is available in the manuscript and supplementary tables

## Acknowledgment

We thank the participants for their cooperation.

We thank the registered nurses, laboratory technicians, nursing students and laboratory technician students from the Arab American University: Ahmad Hodrub, Mohammad Faisal, Wajdi Tùma, Adam Marawà, Sharhabeel Nasrallah, Hisham Zahran and Mohammad Barakat

We thank Medicare labs for providing blood samples of individuals visiting their laboratories.

